# *metaCOVID:* An R-Shiny application for living meta-analyses of COVID-19 trials

**DOI:** 10.1101/2021.09.07.21263207

**Authors:** Theodoros Evrenoglou, Isabelle Boutron, Anna Chaimani

## Abstract

“Living” evidence synthesis is of primary interest for decision-makers to overcome the COVID-19 pandemic. The COVID-NMA provides open-access living meta-analyses assessing different therapeutic and preventive interventions. Data are posted on a platform (https://covid-nma.com/) and analyses are updated every week. However, guideline developers and other stakeholders also need to investigate the data and perform their own analyses. This requires resources, time, statistical expertise, and software knowledge. To assist them, we created the “*metaCOVID”* application which, based on automation processes, facilitates the fast exploration of the data and the conduct of analyses tailored to end-users needs. *metaCOVID* has been created in R and is freely available as an R-Shiny application. The application conducts living meta-analyses for every outcome. Several options are available for subgroup and sensitivity analyses. The results are presented in downloadable forest plots. *metaCOVID* is freely available from https://covid-nma.com/metacovid/ and the source code from https://github.com/TEvrenoglou/metaCovid.

## 1 Introduction

The emergent situation of the COVID-19 pandemic motivated researchers worldwide to rapidly develop and evaluate preventive and therapeutic interventions. In light of an unprecedented explosion of clinical research findings and the uncertainty they are inevitably accompanied with, several initiatives (1–5) were set up to provide ‘living’ evidence on the effects of the different interventions; hence to continuously collect and synthesize all available evidence on COVID-19 treatments, preventive interventions, and vaccines. Such projects aim to serve decision-making by providing global and up-to-date information.

The COVID-NMA platform (https://covid-nma.com/) provides public access to the most up-to-date information with respect to the effects of the different COVID-19 interventions and supports timely clinical decisions and policy-making. Data, analyses, and evidence summaries posted on the platform are updated every week. However, guideline developers and other stakeholders, apart from real-time access to high-quality data, also need to be able to investigate the data and the impact of different characteristics on the results as well as to produce their preferred evidence summaries. For example, different countries have different policies concerning the inclusion or not of preprints when forming COVID-19 guidelines.

To address all these important needs, we developed and made freely available the ‘*metaCOVID’* R-shiny application (https://covid-nma.com/metacovid/). This web-application allows the end-users of the COVID-NMA platform (such as the UK National Institute for Health and Care Excellence and the South African National Department of Health) and other external researchers to directly use the most up-to-date database and perform their preferred meta-analyses in a user-friendly environment. In the present paper, we describe the functionality of the application, the different available options, and the upcoming updates. We aim to provide to current and future users of the application a better understanding of the methods implemented and to illustrate the potential of this tool to produce results tailored to the needs of different stakeholders.

## 2 Implementation

### 2.1 Data

The COVID-NMA database for pharmacologic interventions consists of outcome data, risk of bias assessments, and information on study and population characteristics. We search, on a daily basis to identify new eligible randomized control trials (RCTs) with results and we update the database once a week (1). As of 7 July 2021, the database included 286 RCTs and 213 pharmacologic interventions (monotherapies or drug combinations).

We focus on binary and time-to-event outcomes as well as on both efficacy and safety. The list of available outcomes for synthesis in the application is available in **Appendix Table 1**. Every RCT in our database is accompanied by domain-specific risk of bias assessments (for randomization, missing data, outcome measurement, etc.) as well as by an overall risk of bias assessment based on the Cochrane RoB 2.0 tool (6). In addition, a variety of trial and population characteristics are being extracted from each RCT including the severity of populations, location, type of funding, the presence of conflict of interest, and many other.

### 2.2 Synthesis of findings

The application allows the users to perform a quantitative synthesis of all RCTs that compare the same pair of interventions. For binary outcomes, it calculates automatically the risk ratios (RR) of the RCTs from the 2×2 tables, while for time-to-event outcomes it takes directly the hazard ratios (HR) reported in the trials. The trial-specific effect sizes are synthesized using the inverse-variance method for meta-analysis (i.e. trial weights are proportional to the inverse of their variance). We plan to implement additional synthesis approaches, such as the Mantel-Haenszel method (7).

For the random-effects model that assumes the presence of heterogeneity across the RCTs in a pairwise comparison, we use the restricted maximum likelihood estimator (REML) (8) of the heterogeneity parameter; in case of failure to converge we switch to the Paul-Mantel estimator (9). Given that the identified RCTs appear to be quite heterogeneous we recommend a random-effects meta-analysis as the primary choice. However, the fixed-effect model can also be used either as sensitivity analysis or when there are only 2-3 RCTs that render the estimation of heterogeneity challenging.

Differences among trial results and robustness of meta-analysis findings can be investigated through subgroup and sensitivity analyses respectively. By default, the application groups studies according to the severity of the trial populations and performs meta-analysis for each subgroup as well as for all RCTs together. So far, *metaCOVID* considers only RCTs with hospitalized patients (mild, mixed, or critical) but RCTs on outpatients will also be added. At the same time, a test for subgroup differences (a chi-square test) is provided that suggests whether the summary results from the different subgroups are in statistical agreement (10). Instead of population severity, users may select to perform subgroup analysis according to the location of the RCTs, their type of funding, or their conflict of interest statement.

In every meta-analysis, decisions need to be taken with respect to the handling of missing outcome data as all approaches rely on untestable assumptions. Within the COVID-NMA, we follow a conservative approach by using an intention-to-treat analysis with the assumption that missing outcome data are non-events (1). However, the option of an available case analysis, where participants with missing or incomplete outcome data are excluded from the analysis, is also available and can be used as a sensitivity analysis. We have also implemented an additional sensitivity analysis to investigate the impact of RCTs with a high risk of bias. Specifically, the users can optionally exclude from the analysis all trials assessed at an increased level of risk of overall bias (i.e. high risk and some concerns or high risk only) and check whether there are important changes in the results of the meta-analysis. Finally, there is a lot of discussion on whether preprints should be included in the COVID-19 meta-analyses. Therefore, users can also exclude preprints in a sensitivity analysis and check whether this exclusion impacts the results.

For the synthesis of the results we use the R package *metafor* (11) and we add enhancements to the forest plots (e.g. risk of bias assessments) using self-written functions. The source code of *metaCOVID* is available at https://github.com/TEvrenoglou/metaCovid.

### 2.3 User interface

The application is organized into two domains, analysis options and presentation options, which are described in **Table 1**.

**Table 1:** Summary of the available options in the metaCOVID application.

| Analysis options |  |
| --- | --- |
| Select comparison | Drop-down menu where treatment comparisons are organized according to their type (anti-virals, monoclonal antibodies, etc.) |
| Select outcome | Dropdown menu with all the available outcomes related to the chosen comparison. |
| Meta-analytical model | Provides the choice of whether a random- or a fixed-effect model is going to be used |
| Population severity | Allows the user to focus on a specific type of population based on the severity of the disease |
| Subgroup analysis by | Performs subgroup analysis according to different criteria; the default choice is the severity of the population; pooled results are presented both within and across the subgroups, when there are no subgroups only the overall pooled results are presented. |
| Risk of bias | Allows the exclusion of studies with a high risk of bias either alone or together with those with some concerns for risk of bias; the default choice includes all studies irrespective of their risk of bias |
| Exclude preprints | Allows the exclusion of studies that are not peer-reviewed; the default choice includes all studies irrespective of their publication status |
| Missing outcome data | Allows the user to handle missing data in two different ways. The first choice (the default) is to use all randomized participants and treat missing outcome data as non-events. The second allows for an available case analysis. |
| Presentation options |  |
| Align Risk of bias | Shifts the risk of bias column up and down; this allows the user to better align this column with the rest of the graph. |
| Align Dose | Shifts the dose up and down; this allows the user to better align this column with the rest of the graph. |
| Hide population severity | Allows for hiding the severity of the population reported in each study; this can be useful especially for busy forest plots with many RCTs |
| Hide treatment dose | Allows for hiding the treatment dose reported in each study; this can be useful especially for busy forest plots with many RCTs |

Briefly, the first step for the user is to select the treatment comparison and the outcome of interest. Note that after selecting a treatment comparison, *metaCOVID* scans the database and provides in the list only those outcomes that are available for the chosen treatment comparison. Afterwards, the user can press the button *“Create forest plot”* to obtain the results of the meta-analysis with the default options. The produced forest plot can be downloaded with by pressing *“Download forest plot”*. Alternatively, the user can modify the analysis options (see **Table 1**) and use a different model (e.g. fixed effect) or synthesize a different set of studies within the same comparison based on specific characteristics. This helps exploring how different characteristics of the studies may affect the results. To update the forest plot with the new options, the button *“Create forest plot”* needs to be used again. The user can go back to all the default options by simple pressing *“Reset choices”*.

The user, prior to downloading a forest plot, can improve or change its appearance with the presentation options. This is necessary because the appearance depends on the number of studies in the plot. There is no limit in the number of analyses each user can perform. The procedure starts from the beginning by changing the comparison of interest.

## 3 Use

We illustrate the use of the *metaCOVID* application through a meta-analysis of eleven RCTs that compare Convalescent Plasma versus Standard care or Placebo with respect to the risk of Mortality after 28 days of follow-up. **Figure 1** shows the results of the meta-analysis using the default analysis options and after aligning properly the risk of bias assessments. According to the summary results, the risk of mortality at day 28 is being reduced by 3% with Convalescent plasma in comparison to standard care only. This finding is marginally non-significant. The τ^2^ = 0 suggests the absence of statistical heterogeneity. The graph shows that there is only one RCT with only mild patients while all other RCTs include mixed populations. The test for subgroup differences does not reject the null hypothesis of statistical agreement between the two subgroups.

**Figure 1:**
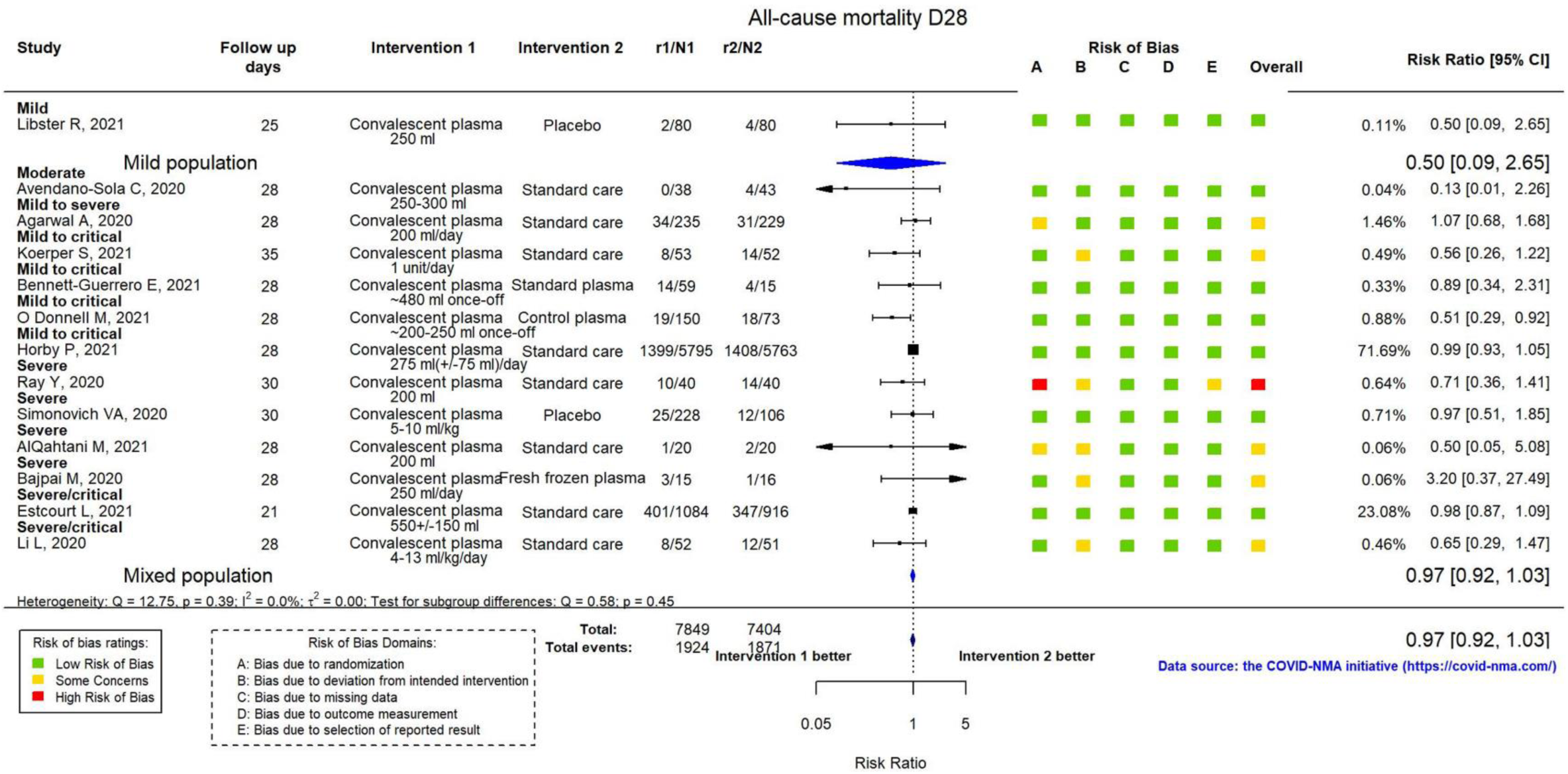
Meta-analysis for the comparison of Convalescent plasma vs Standard care/Placebo for the outcome of mortality at day 28. The studies are separated according to the severity of their population.

We, then, performed a sensitivity analysis excluding studies that have not been published in a peer-reviewed journal yet, but are available as preprints. **Figure 2** suggests that the exclusion of the 6 preprints did not materially affect the results of the meta-analysis. This implies that there is no reason to disregard the RCTs being at the preprint stage.

**Figure 2:**
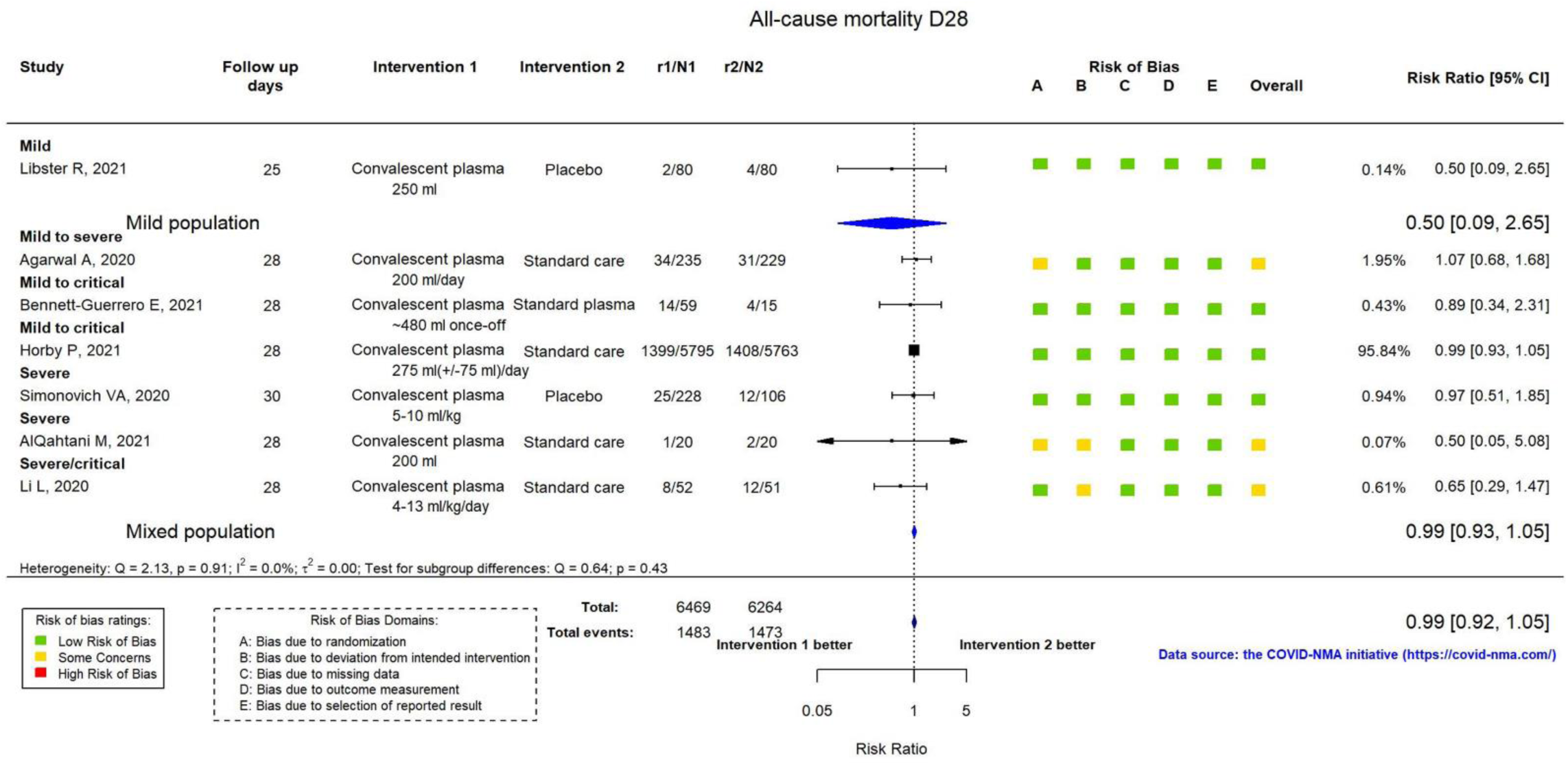
Meta-analysis for the comparison of Convalescent plasma vs Standard care/Placebo for the outcome of mortality at day 28. Only studies published in peer-review journals are included.

Finally, **Figure 3** presents the results from the subgroup analysis based on the presence of conflict of interest. Interestingly, the test for subgroup differences implies that the results from the two groups of trials disagree. This can be seen also from the two diamonds: RCTs without conflict of interest suggest no difference between Convalescent plasma and standard care whereas the two trials with conflict of interest suggest that the drug may substantially reduce the risk of mortality. In other words, it seems that studies having conflict of interest tend to favor the drug. We further see in **Figure 1**, that there is one RCT with high risk of overall bias. Conducting a sensitivity analysis without this RCT or performing an available case analysis did not change materially the results of the meta-analysis (see **Appendix Figure 1** and **Appendix Figure 2**).

**Figure 3:**
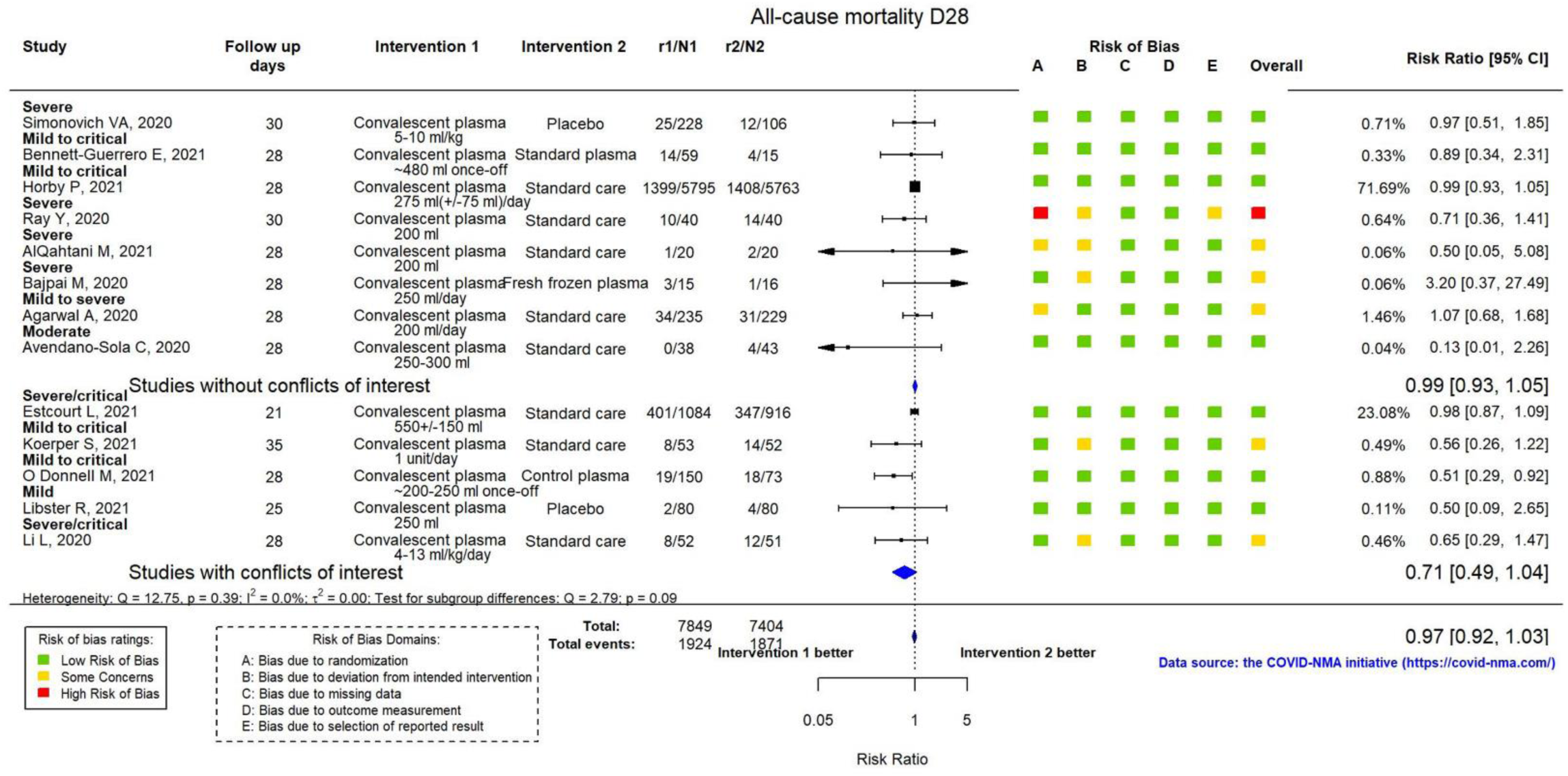
Meta-analysis for the comparison of Convalescent plasma vs Standard care/Placebo for the outcome of mortality at day 28. The studies are separated according to the severity of their conflicts of interest status.

## 4 Discussion

In this paper, we present a new online application, called *metaCOVID*, that provides a user-friendly environment for performing living meta-analyses of COVID-19 trials. Through this application, everyone can use the continuously updated COVID-NMA database to investigate the effects of the different treatments and characteristics that may affect the results. The application offers the opportunity to stakeholders without technical background and software knowledge to easily perform and present complex analyses and data structures.

Since the application is directly connected with the COVID-NMA initiative, which is a ‘living’ project, we also continuously update the *metaCOVID* tool with new functionalities and data. Apart from implementing additional analysis and presentation options (e.g. production of funnel plots, additional meta-analytical models, more variables for subgroup and sensitivity analyses, etc.), one of the key updates planned is to incorporate analyses and data for COVID-19 vaccines.

## Data Availability

All data have been obtained from the COVID-NMA initiative and are available at http://covid-nma.com

## Acknowledgements

The authors would like to thank the whole COVID-NMA consortium and particularly the data extraction team for providing the data used in *metaCOVID* as well as for their useful feedback on the application.

## Appendix

**Appendix Table 1:** Outcomes considered in the COVID-NMA and the metaCOVID application

| Outcome | Timepoint in days (D) | Effect size |
| --- | --- | --- |
| All-cause mortality | D28/D60 or more | Risk ratio |
| Clinical improvement | D28/D60 or more | Risk ratio |
| WHO Clinical Progression Score level 7 or above | D28 / D60 or more | Risk ratio |
| Viral negative conversion | D7 | Risk ratio |
| Incidence of any adverse events | Actual follow-up | Risk ratio |
| Incidence of serious adverse events | Actual follow-up | Risk ratio |
| Time to clinical improvement | Actual follow-up | Hazard ratio |
| Time to death | Actual follow-up | Hazard ratio |
| Time to WHO Clinical Progression Score level 7 or above | Actual follow-up | Hazard ratio |
| Time to viral negative conversion | Actual follow-up | Hazard ratio |

**Appendix Figure 1:**
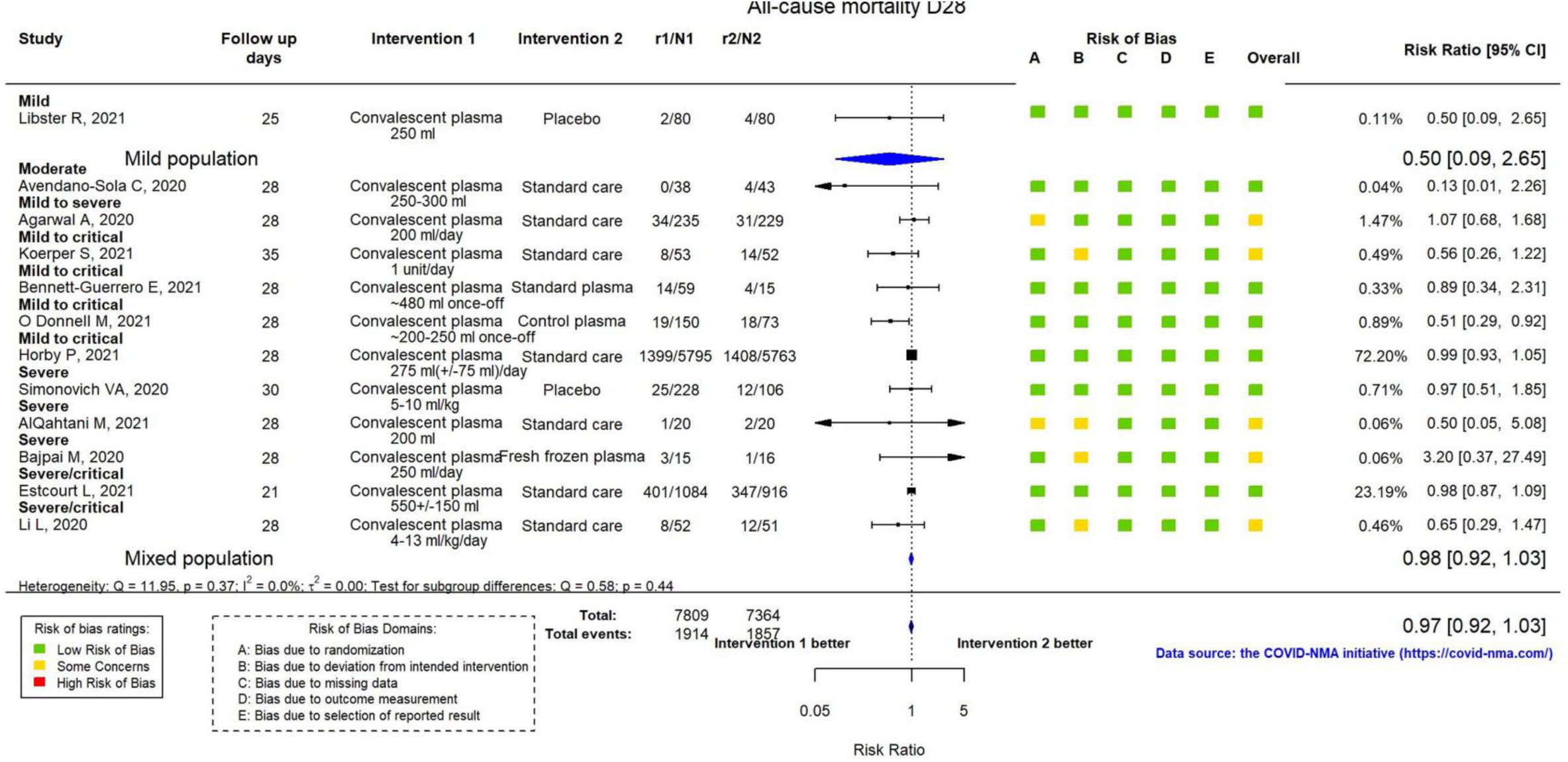
Sensitivity analysis after excluding studies with a high overall risk of bias assessment.

**Appendix Figure 2:**
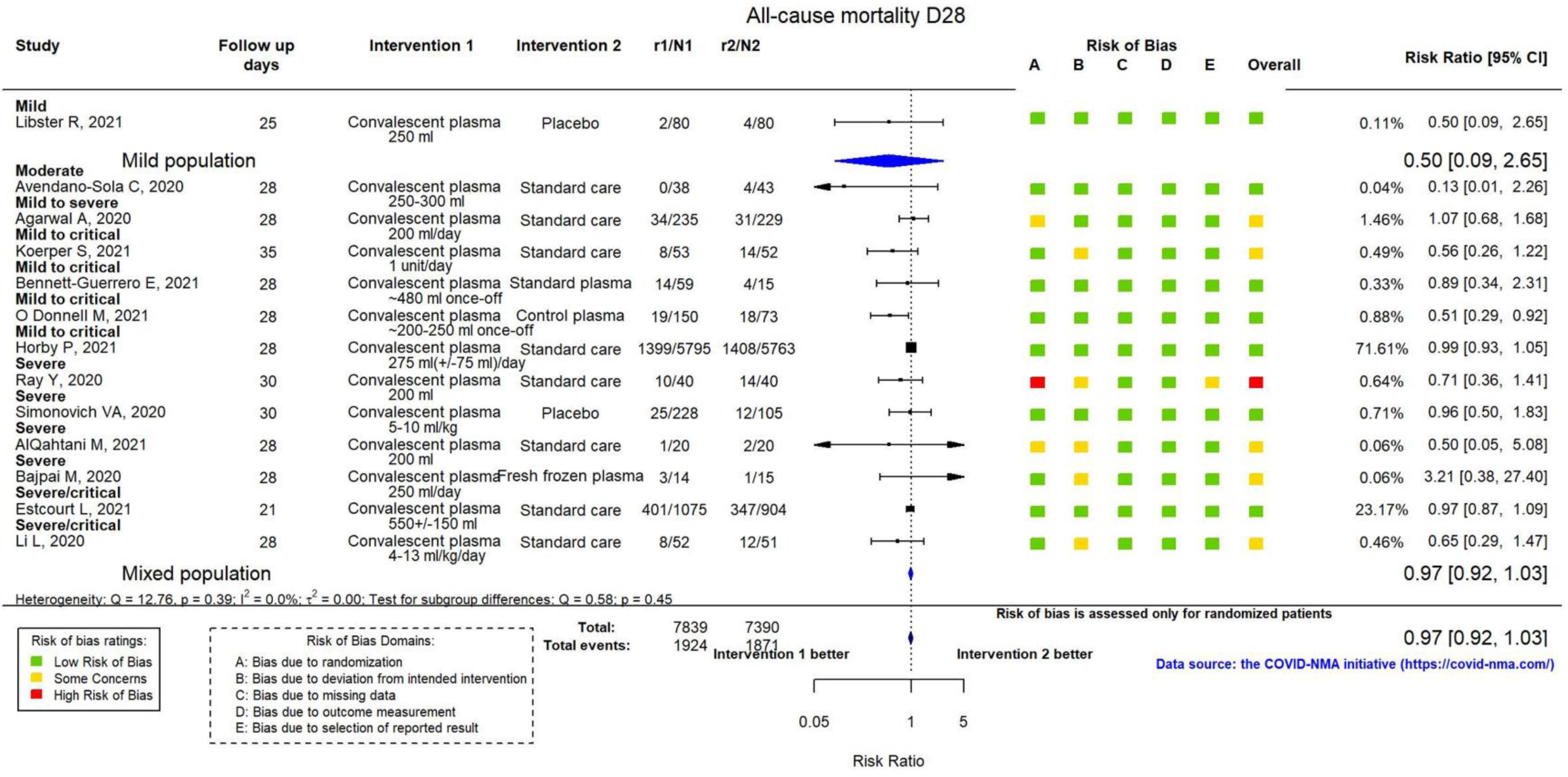
Sensitivity analysis excluding participants with missing or incomplete outcome data (i.e. available case analysis).

